# Evaluating Migraine Trigger Surprisal: Associations with Migraine Activity

**DOI:** 10.1101/2025.04.23.25325821

**Authors:** Dana P. Turner, Twinkle Patel, Emily Caplis, Timothy T. Houle

## Abstract

**Background:** Identifying migraine triggers is a common goal for most individuals with migraine but remains challenging due to the vast number of possible trigger candidates and their fluctuating nature. The *unexpectedness* of daily experiences, quantified through information-theoretic surprisal, may integrate many sources of variation and predict future migraine attacks.

**Objective:** The objective of this study was to evaluate the association between surprisal and future headache attacks.

**Methods:** In this prospective daily diary study, N = 109 individuals with migraine were enrolled, with N = 104 completing twice-daily electronic entries for up to 28 days, yielding 5,176 total diaries. Diary items captured exposure to potential migraine triggers across behavioral, emotional, and environmental domains. For each entry, total surprisal scores were calculated using within-person empirical probability distributions to reflect how atypical each day’s experiences were.

**Results:** Participants experienced a headache on 1,518/5,145 (29.5%) of days with complete diary information. Higher surprisal significantly predicted increased migraine risk within 12 hours (OR = 1.86 [95%CI: 1.12–3.08], p = 0.016) and 24 hours (OR = 2.15 [95%CI: 1.44–3.20], p < 0.001), with stronger effects observed at the longer interval. Notably, the association between current surprisal and migraine onset was moderated by recent surprisal history and exhibited nonlinear properties at 12 hours. Random effects revealed substantial between-person variability in surprisal sensitivity, and individuals with higher baseline headache risk showed attenuated associations.

**Conclusions:** Surprisal offers a novel, individualized measure of trigger unpredictability that is associated with short-term migraine risk. Incorporating surprisal into digital tools may improve personalized prediction and prevention strategies, moving beyond static trigger lists to a dynamic, context-aware model of migraine self-management.

## Introduction

To mitigate the burden of migraine, individuals often strive to identify the triggers, or causes, of their headache attacks.^1–3^ This process relies on covariation assessment, in which one examines how fluctuations in a suspected trigger correspond to variations in migraine occurrence.^4–7^ In practice, this involves evaluating the biopsychosocial factors present before an attack in an attempt to discern meaningful patterns.^8,9^ However, this task is inherently difficult, particularly when the number of potential triggers is vast.^10,11^ Hundreds of factors have been proposed as migraine triggers, ranging from dietary and environmental influences to physiological and psychological stressors, making it nearly impossible for individuals to isolate any single cause with certainty.^2,3,6^

While well-controlled experimental designs could help clarify these associations, they are difficult to implement in everyday life. Instead, people often rely on natural experiments in which both potential causes and migraine outcomes vary freely, making it challenging to determine causal relationships.^8,11,12^ Without a systematic approach, this reliance on observational assessment may lead to inaccurate conclusions, reinforcing beliefs about triggers that may not be scientifically valid.^13^

A robust measurement system capable of evaluating the wide range of possible migraine triggers could provide significant value.^7^ Such a system would need to: (A) accurately quantify the variability in an individual’s exposure to different migraine triggers over time and (B) demonstrate a reliable association between these exposures and the likelihood of near-future migraine attacks. By addressing these challenges, a comprehensive measurement framework could improve migraine prediction and empower individuals with more effective strategies for prevention and management.

We have proposed that headache triggers could be understood not only through their unique physiological mechanisms but also by the degree of surprise they present to individuals.^14^ Using information theory, this work demonstrated that rare or unexpected values of common headache triggers—such as caffeine and alcohol consumption, stress levels, and mood disturbances—were consistently associated with increased headache activity.^15,16^ Summing these individual surprisals into a total trigger surprise score provided stronger discrimination between headache and non-headache days than any single trigger alone.^14^ More recently, research has focused on developing a comprehensive migraine trigger measurement system using surprisal and entropy measures, revealing substantial heterogeneity in exposure patterns both within and between individuals.^17^ A small number of principal components explained most of this variability, suggesting that migraine trigger exposure may be effectively characterized along a few key dimensions. Together, these findings highlight surprisal as a valuable metric for quantifying the unpredictability of trigger exposures and improving headache forecasting.

The current analysis aims to further examine the association between trigger surprisal and future migraine attacks. We hypothesized that the original association observed between surprisal and headache activity^14^ could be replicated in prospectively collected data.^17^ We also illustrate the application of this migraine trigger measurement system by examining the associations of fluctuations in surprisal measurements and near-future migraine activity.

## Methods

This is the second pre-planned primary analysis of these data, with the first analysis being previously published.^17^ After receiving approval from the local Institutional Review Board, this longitudinal daily diary study was conducted between April 2021 and December 2024. Participants were recruited through an online research recruitment platform hosted by our institution, public transportation advertisements, and posted flyers. Interested individuals underwent a telephone screening to determine eligibility. The eligibility criteria have been previously reported but are summarized here for clarity. The criteria for inclusion required having a diagnosis of migraine with or without aura in accordance with the International Classification of Headache Disorders, 3rd Edition (ICHD-3), experiencing between 4 and 14 headache days per month, and being between 18 and 65 years of age. Exclusion criteria included the presence of a secondary headache disorder, chronic daily headache or medication overuse headache, a recent change in headache symptoms within the past six weeks, English proficiency below a sixth-grade level, an unmanaged Axis I psychotic disorder, active substance dependence that could interfere with headache activity and data collection, and pregnancy or planned pregnancy during the study period.

Eligible participants were asked to take part in an enrollment session, either in person or virtually. Prior to participation, individuals completed the informed consent process using the electronic consent feature in REDCap. Following consent, they completed a series of enrollment questionnaires in REDCap (e.g., demographic information, headache characteristics, and the Migraine Disability Assessment [MIDAS]). Participants then received instructions for at-home study procedures, which involved submitting twice-daily diary entries—once in the morning and once in the evening—for 28 days. The diary entries, completed in REDCap, required approximately 5 to 10 minutes. Upon completion of the study procedures, participants took part in a final session, in person or virtually, and completed another series of REDCap questionnaires like those of the enrollment session. Additional questionnaires were completed at study enrollment and completion but are not reported here.

### Daily Diary Items

By design, the twice-daily electronic diaries captured exposure to a wide range of potential migraine triggers, along with measures of headache activity and medication use. The individual items and their variability were described previously.^17^ Given the nature of different trigger constructs, distinct sets of triggers were assessed in the morning (AM) and evening (PM). The AM diary focused on sleep-related behaviors, including duration, quality, awakenings, bedtime, and wake time. It also included questions about late-night meal patterns, weather influences, and mood state using the Profile of Mood States Short Form (POMS-SF).^18^ The PM diary similarly assessed mood state with the POMS-SF but also captured data on commonly reported food and drink triggers, environmental exposures, caffeine and alcohol intake, balance symptoms, meal patterns or missed meals, and weather influences. Additionally, the PM diary included measures of daily stressors from the Daily Stress Inventory (DSI).^19^

### Migraine Trigger Risk Scoring System

To quantify the unexpectedness of an individual’s daily response pattern, we applied the concept of *surprisal* from information theory.^14,16^ Surprisal is defined as the negative logarithm of the probability of an observed exposure (−log[p_exposure_]) and reflects how unlikely that outcome is under a specified probability distribution. In this analysis, empirical probability distributions are constructed separately for each individual using their own observed responses across the study period. For each diary period (i.e., AM and PM each day), the surprisal values of the individual diary items are computed based on these person-specific distributions. The total surprisal is then calculated as the sum of item-level surprisal values for that day and scaled by the number of items to yield an average surprisal per item. This scaling allows for missing items to be ignored and for AM diaries to be scaled similarly to the PM diaries despite the fact that the latter has more items (see:^17^). This approach provides a within-person measure of how atypical a given day’s responses are relative to that individual’s own behavioral patterns.

In our recent work,^17^ the calculation of total surprisal incorporated two components: surprisal based on the magnitude of each measurement and surprisal associated with the change from the preceding value. This required estimating and summing surprisal values from two within-person empirical distributions: the distribution of observed values and the distribution of within-person changes (i.e., X_t_ − X_t−1_) for each variable. In the current analysis, we simplify this approach by omitting the change-based component of surprisal. Instead, we include a lagged total surprisal term (Surprisal_t−1_) in the statistical models to account for short-term fluctuations in surprisal over time.

### Outcomes

The primary outcomes for this analysis are two binary variables indicating the occurrence of a headache attack within a specified future time window. Specifically, we define a dichotomous outcome representing whether a headache attack occurs within the subsequent 12 hours (0 = no attack, 1 = attack) and a second outcome representing whether an attack occurs within the subsequent 24 hours (0 = no attack, 1 = attack). An attack was defined as any self-reported headache with pain > 0 and any pattern of secondary symptoms (e.g., photophobia, phonophobia). These outcomes are aligned to each diary entry time point (e.g., 12 hours from the AM entry) and allow us to evaluate short-term prospective associations between daily surprisal and the onset of headache attacks over two clinically relevant temporal horizons.

### Statistical Power Considerations

The originally planned sample size of N = 200 provided for 95%CI precision of +-4.16% for an observed proportion of 10%. However, the COVID-19 pandemic limited the number of participant enrollments. Our institution mandated a pause of research and hiring of new staff for a long time period. Further, potential participants who were concerned about virus transmission were cautious about enrolling when research activity opened again. The final enrollment number was N = 109. This is a sufficient number because these hypotheses can be meaningfully evaluated with a smaller than intended sample.

### Statistical Analyses

All statistical analyses were conducted using R version 4.4.1 and RStudio. Descriptive statistics were used to summarize sample characteristics, diary adherence, and the distribution of surprisal scores across participants and time points. Means and standard deviations were computed for continuous variables, while frequencies and percentages were used for categorical variables. To examine the association between daily surprisal scores and the likelihood of subsequent migraine attacks, generalized linear mixed-effects models (GLMMs) were fitted using the ‘lme4’ package. These models accounted for the nested structure of the data (i.e., repeated diary entries within individuals) by including a random intercept for participant. In the primary analyses, the fixed effects included the total surprisal score at time *t*, with random slopes for surprisal to capture inter-individual variability in the surprisal-headache association. Models were estimated separately for two binary outcomes using a binomial likelihood and logit link: the occurrence of a migraine attack within 12 hours and within 24 hours of a diary entry. To explore temporal dependencies, each model included the lagged surprisal score from the previous diary entry (Surprisal_t−1_) as a fixed effect. In secondary models, we tested for nonlinear and interactive effects by including a squared term for current surprisal and its interaction with the lagged surprisal score. Model fit was evaluated using likelihood ratio tests and information criteria (AIC, BIC), and model assumptions were verified through residual diagnostics. Effect estimates are reported as odds ratios (ORs) with 95% confidence intervals (CIs). Missing data were ignored for the analysis, with all models conducted on complete cases. A two-tailed p-value less than 0.05 was considered statistically significant for all hypothesis tests. Intraclass correlation coefficients (ICCs) and variance components were calculated to quantify between-person differences in baseline risk and in the strength of the surprisal effect. In sensitivity analyses, models were adjusted for each participant’s average surprisal score to better isolate within-person effects. All models were fit using maximum likelihood estimation with the ‘glmer’ function.

## Results

The sample characteristics have been previously reported.^17^ Briefly, the study enrolled 109 participants (median age: 35 years [26.0, 46.0]), the majority of whom were female (102, 93.5%) and White (91, 83.5%). Of the N = 109 individuals with migraine who were enrolled, N = 104 completed twice-daily electronic entries for up to 28 days, yielding 5,176 total diaries. All participants had migraine, with a median headache frequency of 8 [5.0, 12.0] days per month and a median intensity of 7/10 [5.5, 8.0]. Most had headaches that were unilateral (79, 72.5%) and pulsating (57, 52.3%), with common symptoms including photophobia (105, 96.3%), phonophobia (95, 89.6%), and nausea/vomiting (99, 90.8%). The median MIDAS score of 24 [13.0, 35.5] indicated moderate to severe disability.

### Distribution of Surprisal Scores

Table 1 presents the pooled distribution of surprisal scores for all participant days in relation to future headache attacks across different diary entries and summarizes the mean pooled surprisal scores (± SD) by diary type (AM vs. PM), current headache presence, and future headache attacks at 12- and 24-hour intervals. Slightly higher surprisal scores were observed in the presence of a current headache (0.06 to 0.10 bits) and when a future headache would develop (0.01 to 0.06 bits), with the highest values occurring in the PM diary entries (e.g., PM, current headache, future headache in 12 hours: 0.74 ± 0.24). These subtle differences indicate that when pooled across individuals, the surprisal scores might allow only modest discrimination across headache days.

**Table 1.** Daily Characteristics of Scaled Surprisal Scores.

| Diary Type | Current Headache | Future Headache Attack 12 hours |  | Future Headache Attack 24 hours |  |
| --- | --- | --- | --- | --- | --- |
|  |  | No | Yes | No | Yes |
| AM | No | 0.53 (0.18) | 0.56 (0.19) | 0.53 (0.18) | 0.55 (0.18) |
|  | Yes | 0.61 (0.21) | 0.62 (0.19) | 0.61 (0.22) | 0.62 (0.18) |
| PM | No | 0.62 (0.24) | 0.64 (0.24) | 0.60 (0.23) | 0.65 (0.25) |
|  | Yes | 0.70 (0.26) | 0.74 (0.24) | 0.67 (0.25) | 0.73 (0.25) |

### Association of Surprisal Total Score and Future Headache Activity

Participants experienced a headache on 1,518/5,145 (29.5%) of days with complete diary information. The surprisal total score is a significant predictor of both 12-hour and 24-hour headache attack risk, with odds ratios of 1.86 (95%CI: 1.12–3.08, *p* = 0.016) and 2.15 (95%CI: 1.44–3.20, *p* < 0.001), respectively. These findings suggest that for each average bit increase in surprisal, the likelihood of a future headache also rises, with a stronger effect at 24 hours than at 12 hours. Table 2 displays the models used for 12 and 24 hours, and Figure 1 displays this association along with the distribution of individual surprisal slopes for each outcome.

**Table 2.** Association of Surprisal Total Score and Future Headache Attack.

| Predictors | Future Attack (12 Hours) |  |  | Future Attack (24 Hours) |  |  |
| --- | --- | --- | --- | --- | --- | --- |
|  | OR | CI | p | OR | CI | p |
| (Intercept) | 0.68 | 0.49 – 0.96 | 0.027 | 0.53 | 0.39 – 0.71 | <0.001 |
| Current headache (Yes) | 1.88 | 1.61 – 2.20 | <0.001 | 1.64 | 1.43 – 1.89 | <0.001 |
| Diary type (PM) | 0.47 | 0.41 – 0.55 | <0.001 | 0.82 | 0.72 – 0.93 | 0.002 |
| Surprisal | 1.86 | 1.12 – 3.08 | 0.016 | 2.15 | 1.44 – 3.20 | <0.001 |
| **Random Effects** |  |  |  |  |  |  |
| $\sigma^2$ | 3.29 | | | 3.29 | | |
| $\tau_{00}$ | 0.81 | | | 0.54 | | |
| $\tau_{11}$ | 2.00 | | | 0.31 | | |
| $\rho_{01}$ | -0.81 | | | -0.50 | | |
| ICC | 0.11 |  |  | 0.11 |  |  |
| N | 103 |  |  | 104 |  |  |
| Observations | 4530 |  |  | 4947 |  |  |
| Marginal $R^2$ / | 0.054 / | | | 0.024 / | | |
| Conditional $R^2$ | 0.157 | | | 0.136 | | |

**Figure 1.**
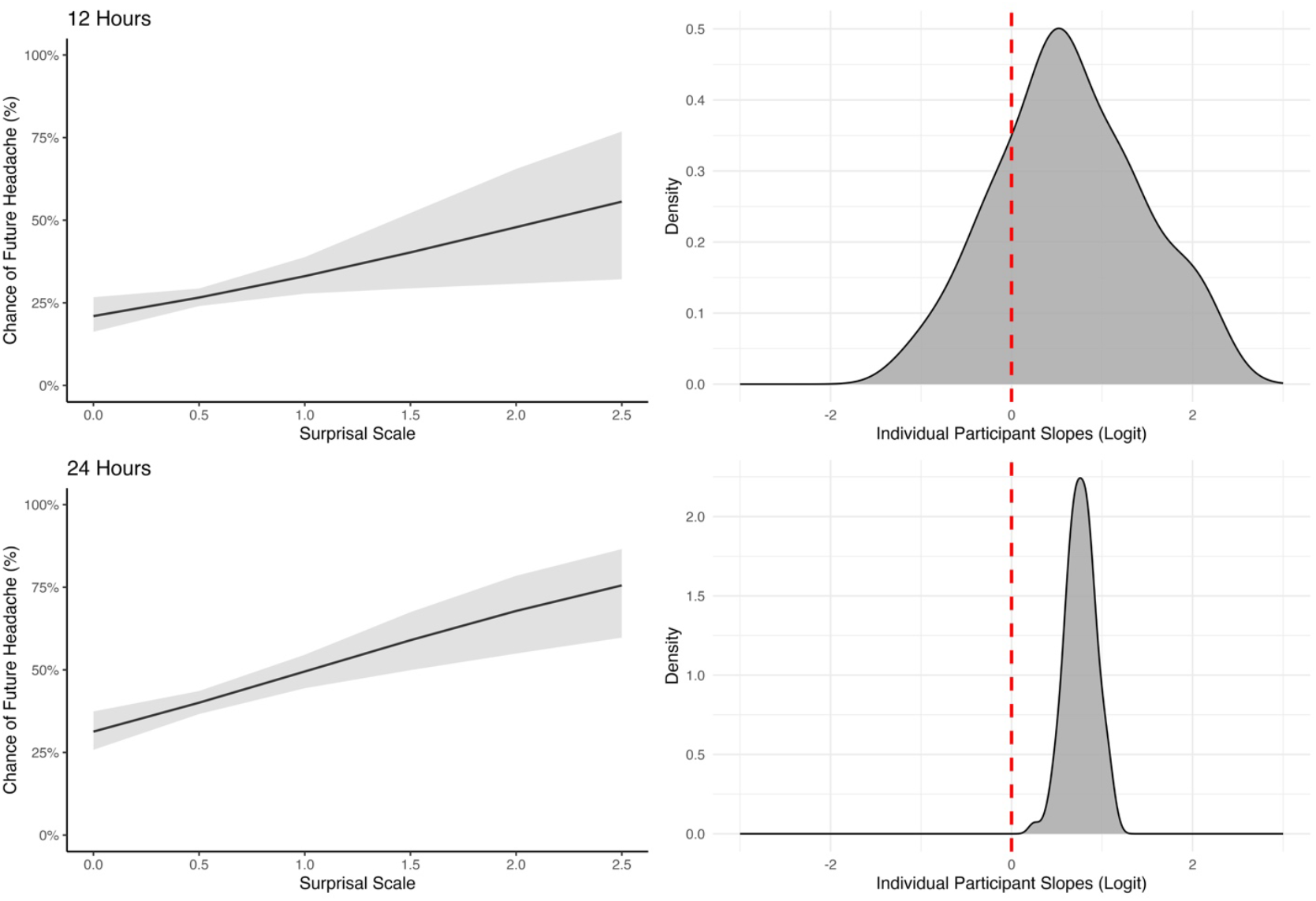
Model-predicted probability of future headache onset as a function of surprisal, shown separately for 12-hour (top left) and 24-hour (bottom left) prediction windows. Shaded areas represent 95% confidence intervals. The right panels display the distributions of individual surprisal slopes from the mixed-effects models on the logit scale, highlighting variability in person-level associations. Red dashed lines denote zero slope.

The random variance components indicate substantial between-person variability in how surprisal influences headache risk. Specifically, the random slope variance (τ_11_) for surprisal is 2.00 at 12 hours but notably lower at 0.31 for 24 hours, suggesting that individuals differ more in their immediate response to surprisal than in its longer-term effects (See also Figure 1). The negative correlation between random intercept and surprisal slope (ρ_01_ = −0.81 for 12 hours, −0.50 for 24 hours) further implies that individuals with higher baseline headache risk exhibit a weaker association between surprisal and attack likelihood. The intraclass correlation coefficient (ICC = 0.11 for both models) indicates that 11% of the variance is attributable to individual differences, reinforcing the importance of person-level variability in headache prediction. In a sensitivity model, each individual’s average surprisal score was further adjusted to better consider between-person variation. In these models, the association between total surprisal and attack risk was somewhat attenuated for 12 hours, OR 1.56 (95%CI: 1.01 to 2.40), p = 0.044, and 24 hours, OR 1.88 (95%CI: 1.27 to 2.79), p = 0.002. Overall, these results highlight surprisal as a key predictor of headache onset, with individual differences potentially playing a critical role in shaping this association over time.

### Nonlinear and Contextual Associations of Total Score and Future Headache Activity

For the 12-hour model predicting the likelihood of a future headache episode, results indicated a significant interaction between the current surprisal score and its lagged value, along with evidence supporting a nonlinear (quadratic) relationship. The best-fitting model included both linear and squared surprisal terms, the lagged surprisal term, and their interactions, as determined through likelihood ratio tests and model selection criteria (AIC, BIC). As shown in the left panel of Figure 2 and Table 3, higher current surprisal was associated with an increased probability of a headache within 12 hours, but this relationship varied depending on prior surprisal. When prior surprisal was low (lag = 0.35), the effect of current surprisal was more strongly positive and nonlinear, with headache probability increasing steeply beyond a surprisal score of approximately 1. In contrast, at higher lag values (e.g., 0.9), the association flattened or even reversed. The interaction between the current surprisal squared and its lagged value was statistically significant (OR = 0.02, 95%CI [0.00–0.43], *p* = 0.014), highlighting how recent context can modulate the impact of momentary experiential dynamics.

**Table 3.** Contextual Associations of Surprisal Total Score and Future Headache Attack.

| Predictors | Future Attack (12 Hours) |  |  | Future Attack (24 Hours) |  |  |
| --- | --- | --- | --- | --- | --- | --- |
| | OR | CI | $p$ | OR | CI | $p$ |
| (Intercept) | 0.75 | 0.25 – 2.31 | 0.620 | 0.28 | 0.15 – 0.51 | **<0.001** |
| Current headache (Yes) | 1.86 | 1.59 – 2.18 | **<0.001** | 1.63 | 1.41 – 1.89 | **<0.001** |
| Diary type (PM) | 0.51 | 0.44 – 0.59 | **<0.001** | 0.80 | 0.70 – 0.92 | **0.002** |
| Surprisal | 0.11 | 0.00 – 2.84 | 0.185 | 5.95 | 2.54 – 13.95 | **<0.001** |
| Surprisal<sup>2</sup> | 10.66 | 1.12 – 101.15 | **0.039** |  |  |  |
| Surprisal<sub>t-1</sub> | 0.64 | 0.09 – 4.37 | 0.647 | 3.54 | 1.52 – 8.22 | **0.003** |
| Surprisal × Surprisal<sub>t-1</sub> | 137.48 | 0.82 – 22922 | 0.059 | 0.20 | 0.07 – 0.59 | **0.004** |
| Surprisal<sup>2</sup> × Surprisal<sub>t-1</sub> | 0.02 | 0.00 – 0.43 | **0.014** |  |  |  |
| **Random Effects** |  |  |  |  |  |  |
| $\sigma^2$ | 3.29 | | | 3.29 | | |
| $\tau_{00}$ | 0.33 | | | 0.43 | | |
| ICC | 0.09 |  |  | 0.12 |  |  |
| N | 103 |  |  | 103 |  |  |
| Observations | 4440 |  |  | 4440 |  |  |
| Marginal R<sup>2</sup> / | 0.080 / |  |  | 0.031 / |  |  |
| Conditional R<sup>2</sup> | 0.164 |  |  | 0.144 |  |  |

**Figure 2.**
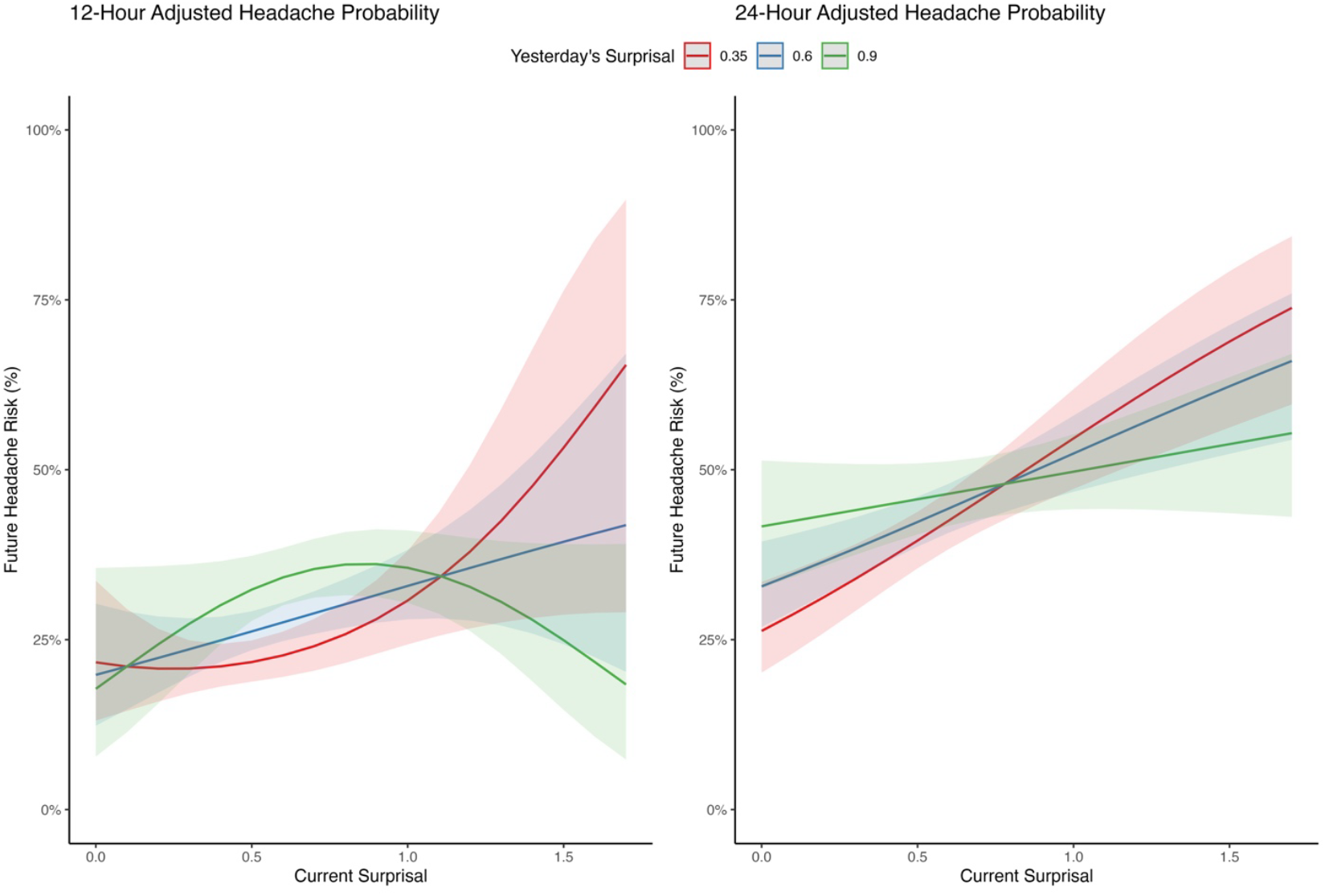
Model-predicted probability of future headache occurrence as a function of current surprisal, conditioned on different levels of lagged surprisal from the previous day (0.35, 0.6, and 0.9). Shaded areas represent 95% confidence intervals. The left panel shows predictions for the 12-hour window, and the right panel shows predictions for the 24-hour window, illustrating how recent surprisal history modulates the impact of current trigger states on headache risk.

In contrast, the 24-hour model demonstrated a simpler, largely linear pattern in the relationship between surprisal and headache probability. This model did not benefit from including a quadratic term, though it retained the lagged surprisal predictor and its interaction with the current surprisal score. As depicted in the right panel of Figure 2 and Table 3, higher current surprisal predicted a greater probability of headache 24 hours later, but the strength of this relationship decreased as the lagged surprisal increased. Specifically, at low lag values (lag = 0.35), surprisal had a steep positive slope, with the odds of headache increasing nearly sixfold (OR = 5.95, 95%CI [2.54–13.95], *p* < 0.001). However, at higher lag values, this effect was attenuated, consistent with a significant negative interaction between current surprisal and its lagged counterpart (OR = 0.20, 95%CI [0.07–0.59], *p* = 0.004). These findings suggest that while current surprisal is a robust predictor of subsequent headache activity, its influence is temporally dynamic, moderated by recent history.

## Discussion

This study evaluated the relationship between daily surprisal scores—quantifying the unexpectedness of daily experiences—and the subsequent occurrence of migraine attacks within 12- and 24-hour windows. Using intensive longitudinal data and person-specific statistical modeling, we found that higher surprisal scores were meaningfully associated with an increased risk of future headache attacks. These results support our initial hypothesis and replicate earlier findings^14^ in a new, prospectively collected dataset, underscoring surprisal as a valuable metric for forecasting migraine risk.

The results provide compelling evidence that the degree to which an individual’s experiences deviate from their usual patterns, measured by information-theoretic surprisal, can meaningfully predict migraine activity. For each unit increase in average surprisal, the odds of a migraine attack rose substantially, with a stronger effect observed over the 24-hour period than the 12-hour time window. This suggests that the influence of daily experiences on migraine onset is not immediate in all cases, exhibits a considerable delay in effect, and may accumulate or evolve over time. The finding that prior surprisal moderates this relationship adds further nuance: the predictive strength of current surprisal was attenuated, or even reversed, when recent surprisal levels were already elevated, indicating a possible contextual adaptation or regulatory effect.

A key strength of this study lies in its examination of individual differences. The random slope models revealed considerable variability in how surprisal influenced headache risk across participants. Some individuals appeared more sensitive to fluctuations in daily life, while others showed little to no association between surprisal and migraine. Interestingly, those with higher baseline headache risk tended to show weaker associations with surprisal, as indicated by the strong negative correlation between random intercepts and slopes. This pattern raises the possibility that migraine activity in these individuals may be more influenced by stable factors (e.g., neurobiological predisposition) than by daily behavioral or environmental changes. Such findings have important implications for the development of personalized headache management tools.

From a clinical perspective, these findings suggest that surprisal may be a useful construct for advancing migraine self-management. Current trigger-tracking approaches often emphasize binary relationships between exposures and outcomes, potentially overlooking the dynamic and context-dependent nature of real-life experiences. By contrast, surprisal offers an integrative, continuous, person-centered measure that reflects how unusual a given day is for a specific individual. Embedding surprisal scores into digital headache diaries or wearable technologies could enable real-time risk forecasting and provide users with actionable insights, such as identifying patterns of instability or identifying days when risk is particularly elevated.^9,20–22^ Novel interventions might focus not on avoiding specific triggers (see:^23–25^) but on incorporating behavioral regularity or emotional regulation to control the experience of surprisal itself.

Our results align with and extend prior work demonstrating the value of information-theoretic approaches in migraine research.^14,15,17^ Earlier studies showed that surprisal scores derived from diary data could discriminate headache days from non-headache days and that these scores captured meaningful within-person variation in trigger exposures.^14^ In addition to replicating past findings, the current study adds several critical dimensions by showing that surprisal is not only associated with current headache status but also predicts different aspects of future risk (i.e., 12 versus 24 hours). Additionally, the use of lagged surprisal values and different times of day (i.e., AM and PM diaries) highlights the importance of context in interpreting the impact of daily exposures—a feature that has received little attention in our prior migraine forecasting models.^9,20^

This study has several particular strengths. The prospective, real-time design with twice-daily entries allowed for fine-grained measurement of behavioral and environmental exposures. The use of person-centered empirical probability distributions ensured that surprisal estimates were tailored to individual patterns rather than relying on group averages. High adherence rates and a rich dataset of nearly 6,000 diary entries further enhanced the robustness of the analyses.

However, limitations should be acknowledged. The final sample size, while sufficient for the modeling approach, was smaller than initially planned due to COVID-19-related disruptions. The sample was predominantly White and female, which may limit generalizability. Additionally, the current analysis focused solely on magnitude-based surprisal, omitting the change-based component used in earlier work. While lagged surprisal helped account for short-term fluctuations, future models may benefit from further consideration of integrating both components. We also did not include potential covariates such as acute medication use, which may interact with surprisal to influence migraine risk.

Future research should develop methods to estimate surprisal prospectively and consider ways to make the scoring method less burdensome. For example, completing hundreds of different items daily samples a host of behavioral and environmental factors from which to estimate a total surprisal score, but the time required could be prohibitive for some individuals, especially over extended lengths of time. Future work should also explore the integration of surprisal into real-time digital platforms capable of dynamically predicting migraine onset. Investigating neurobiological, psychological, or lifestyle correlates of high surprisal sensitivity may yield additional insight into personalized prevention strategies. Extending this work to larger, more clinically heterogeneous samples and across longer observation periods will be important for validating the generalizability and clinical utility of surprisal-based models.

In conclusion, this study supports using a surprisal scoring system as a dynamic, individualized metric capable of predicting short-term migraine risk. The findings underscore the value of a person-centered, information-theoretic approach to understanding migraine triggers, one that moves beyond static lists of potential causes to account for the unpredictable and context-sensitive nature of daily life. Incorporating surprisal into migraine forecasting tools could provide individuals with a more effective, personalized strategy for managing headache risk.

## Data Availability

Data may be available upon reasonable request to the authors.

## References

1. Seng EK, Martin PR, Houle TT. Lifestyle factors and migraine. Lancet Neurol. 2022;21:911–21.

2. Pellegrino ABW, Davis-Martin RE, Houle TT, Turner DP, Smitherman TA. Perceived triggers of primary headache disorders: A meta-analysis. Cephalalgia. 2018;38:1188–98.

3. Pavlovic JM, Buse DC, Sollars CM, Haut S, Lipton RB. Trigger factors and premonitory features of migraine attacks: Summary of studies. Headache. 2014;54:1670–9.

4. Turner DP, Leffert LR, Houle TT. Appraisal of headache trigger patterns using calendars. Headache. 2019;60:370–81.

5. Vives-Mestres M, Casanova A, Buse DC, Donoghue S, Houle TT, Lipton RB, et al. Patterns of perceived stress throughout the migraine cycle: A longitudinal cohort study using daily prospective diary data. Headache. 2021;61:90–102.

6. Marmura MJ. Triggers, protectors, and predictors in episodic migraine. Curr Pain Headache Rep. 2018;22:81. doi: 10.1007/s11916-018-0734-0.

7. Turner DP. Assessing headache triggers: A practical guide for applied research and clinical management. Cham, Switzerland: Springer Nature. 2021.

8. Turner DP, Houle TT. Learning headache triggers through experience: A laboratory study. Headache. 2023;63:721–9.

9. Houle TT, Turner DP, Golding AN, Porter JAH, Martin VT, Penzien DB, et al. Forecasting individual headache attacks using perceived stress: Development of a multivariable prediction model for persons with episodic migraine. Headache. 2017;57:1041–50.

10. Lipton RB, Penzien DB, Turner DP, Smitherman TA, Houle TT. Methodological issues in studying rates and predictors of migraine progression and remission. Headache. 2013;53:930–4.

11. Turner DP, Smitherman TA, Martin VT, Penzien DB, Houle TT. Causality and headache triggers. Headache. 2013;53:628–35.

12. Turner DP, Houle TT. Influences on headache trigger beliefs and perceptions. Cephalalgia. 2018;38:1545–53.

13. Houle TT, Turner DP. Natural experimentation is a challenging method for identifying headache triggers. Headache. 2013;53:636–43.

14. Turner DP, Lebowitz AD, Chtay I, Houle TT. Headache triggers as surprise. Headache. 2019;59:495–508.

15. Turner DP, Caplis E, Bertsch J, Houle TT. Information theory and headache triggers. Headache. 2023;63:899–907.

16. Shannon CE. A mathematical theory of communication. The Bell System Technical Journal. 1948;27:379–423.

17. Turner DP, Caplis E, Patel T, Houle TT. Development of a migraine trigger measurement system using surprisal. MedRxiv. 2025. medRxiv doi: 10.1101/2025.02.27.25322488

18. Shacham S. A shortened version of the Profile of Mood States. J Pers Assess. 1983;47:305–6.

19. Brantley PJ, Waggoner CD, Jones GN, Rappaport NB. A Daily Stress Inventory: Development, reliability, and validity. J Behav Med. 1987;10:61–74.

20. Houle TT, Deng H, Tegeler CH, Turner DP. Continuous updating of individual headache forecasting models using Bayesian methods. Headache. 2021;61:1264–73.

21. Turner DP, Leffert LR, Houle TT. The treatment implications of forecasting headache. Pain Manag. 2020;10:349–352.

22. Turner DP, Lebowitz AD, Chtay I, Houle TT. Forecasting migraine attacks and the utility of identifying triggers. Curr Pain Headache Rep. 2018;22:62. doi: 10.1007/s11916-018-0715-3.

23. Martin PR, Reece J, Callan M, MacLeod C, Kaur A, Gregg K, et al. Behavioral management of the triggers of recurrent headache: A randomized controlled trial. Behav Res Ther. 2014;61:1–11.

24. Martin PR. Headache triggers: To avoid or not to avoid, that is the question. Psychol Health. 2000;15:801–9.

25. Martin PR. Managing headache triggers: think “coping” not “avoidance.” Cephalalgia. 2010;30:634–7.

